# Impact of proteogenomic evidence on clinical success

**DOI:** 10.64898/2026.02.23.26346731

**Authors:** Mohd Anisul Karim, Abhay Hukku, Bruno Ariano, Emily Holzinger, Yakov Tsepilov, James Hayhurst, Annalisa Buniello, Ellen M. McDonagh, Stephane E Castel, Matthew R. Nelson, Joseph Maranville, Laura Yerges-Armstrong, Maya Ghoussaini

## Abstract

We assessed the impact of plasma protein quantitative trait loci (pQTL) on genetically supported therapeutic hypotheses. We show that pQTL-supported target-indication pairs were 4.4 times more likely to advance from Phase I to launch, compared to a 2.6-fold increase observed with other human genetic evidence. Moreover, pQTL-based enrichment was prominent in druggable protein families which had limited enrichment from other genetic evidence sources.

## Main

Suboptimal therapeutic efficacy is a key reason for the failure of a drug for its selected disease indication. Several reports have shown that drug targets matched to disease indications via human genetic evidence have at least a two-fold higher probability of clinical success^1–3^. Genome-wide association studies (GWAS) of diseases are a major source of human genetic evidence but genome-wide association signals do not directly implicate causal genes, as lead variants may lie in intergenic regions or in linkage disequilibrium with variants affecting several nearby genes, limiting the ability of GWAS alone to assign causal genes with confidence^4^. An approach to assign causal genes with higher confidence is to evaluate whether the disease-associated genetic signal shares causal variants with (that is, colocalizes with) genetic signals from intermediate molecular traits such as plasma proteins. Indeed the probability of clinical success of drug programs rises with increasing confidence that a particular gene is causal for disease^1^. Prior studies have mostly examined the enrichment of pQTLs in druggable targets to uncover new therapeutic opportunities^5,6^. A more recent study showed clinical success enrichment with cis-QTLs broadly (pQTL & eQTL)^7^, and a composite genetic priority score integrating eight genetic features, including pQTL overlap, was associated with drug indication status and clinical phase progression^8^. However, neither study isolated the specific contribution of pQTL evidence to clinical success, and it remained unclear whether broader pQTL-based MR and colocalization support adds value beyond standard human genetic evidence.

Here, we attempt to expand on these studies including our own previous work^9^ by integrating eight publicly available plasma proteogenomic datasets (**Supplementary Table 1**) with a broad collection of over 8000 complex traits from GWAS Catalog, Pan-UK Biobank, and FinnGen (**Supplementary Table 2**) and a recent large curated set of 29,476 drug target–indication (T–I) pairs from Citeline Pharmaprojects^1^. To assign genes to indications matching GWAS traits, Minikel et al^1^ derived a locus-to-gene (L2G) share which is the proportion of total L2G score assigned to a gene from among all possible locus-gene-trait triplets in Open Targets Genetics (version 8). The L2G score is derived from a trained supervised learning model that integrates multiple lines of evidence including genomic distance, chromatin interaction data, and functional annotations, especially cis-QTLs including cis-QTLs from GTEx, eQTLGen and the eQTL Catalogue, and cis-pQTLs from a single 3,301-participant plasma proteomic study^10^, to predict causal genes at GWAS loci^4^. Notably, pQTL evidence contributed to fewer than 1% of loci in that framework^4^. Because our analyses are not limited to cis-pQTLs from a single study, we investigated whether having broader pQTL-support can potentially complement L2G-based therapeutic hypotheses.

To ensure comparability of pQTL-supported genetic hypotheses with other sources of human genetic evidence, we used the same MeSH (MeSH similarity score > 0.8) and L2G thresholds (L2G share > 0.5) as Minikel et al in our main analysis, but varied the L2G thresholds by +0.25 increments to evaluate the added value of pQTL-supported hypotheses. To generate pQTL-supported therapeutic hypotheses, we first performed a large-scale systematic MR - 47.2M tests - using genome-wide significant genetic instruments associated with plasma protein levels and ∼8000 GWAS traits. We merged these significant MR target-trait associations with gene– indication pairs from Minikel et al’s T-I dataset and retained pairs where our MR outcomes matched their known drug indications (MeSH similarity ≥ 0.8). For the enrichment analysis comparing pQTL-supported versus unsupported hypotheses, we further restricted to target-indication pairs with both MeSH similarity ≥ 0.8 and L2G share ≥ 0.5 within the background of proteomically measured genes, yielding 49 pQTL-supported target-indication pairs at Phase I used for relative success calculation (**Supplementary Figure S1)**.

In our enrichment analyses, we found that pQTL-supported target-indication pairs showed a substantial enrichment for clinical success (Phase I to Launch), with a relative success (RS) of 4.44 (95% CI: 3.27, 6.02); this is broadly comparable to OMIM-based evidence (RS: 3.67, 95% CI: 3.07, 4.37) (**Fig. 1a, Supplementary Table 4**). Notably, T–I pairs with both pQTL support and an L2G share of ≥0.75 achieved an RS of 5.13 (95% CI: 3.73, 7.06), approximately double the RS for L2G ≥0.75 alone (RS of 2.60; 95% CI: 2.17, 3.13), suggesting added value of pQTLs to prioritise therapeutic hypotheses. Evaluated without any L2G requirement, pQTL support was associated with a relative success of 3.37 (95% CI: 2.46, 4.62), increasing the number of supported target–indication pairs from 88 to 128 (**Supplementary Table 4**). pQTL support also retained predictive value where rare-variant and gene-burden evidence were absent, with a relative success of 4.52 (95% CI: 3.13, 6.52) among pairs carrying neither OMIM nor Genebass support, which accounted for 71% of pQTL-supported pairs; these sources are therefore largely non-overlapping and complementary in coverage, and pQTL support added little among pairs already carrying OMIM evidence (**Supplementary Table 19**). Targets assayed on the proteomics platforms but without pQTL support were launched at 10.6% from Phase I, below the 12.7% observed for targets not assayed on any platform (relative success 0.83; 95% CI: 0.76, 0.92), indicating that panel membership does not inflate the comparator (**Supplementary Figure S4**).

**Figure 1.**
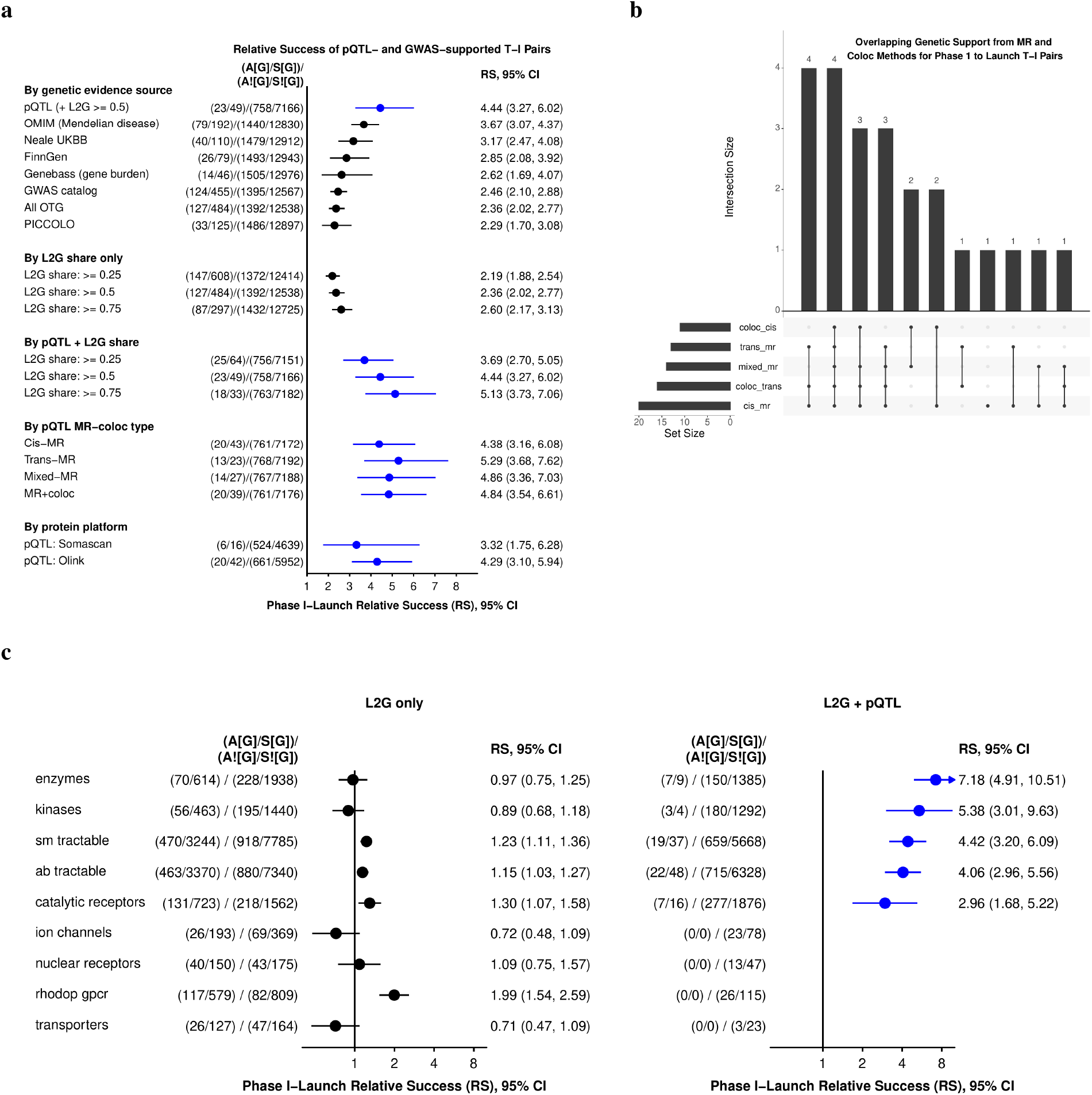
Enrichment analyses. **a**. Phase I–Launch relative success (RS) of target–indication (T–I) pairs, comparing pQTL-based evidence (blue) with non-pQTL evidence (black) by genetic evidence source, L2G share alone, L2G share plus pQTL evidence, MR-coloc type, and protein assay platform. pQTL-based RS used T–I pairs with L2G share > 0.5 and Bonferroni-significant pQTL support (p < 1.06 × 10^−9^). Non-pQTL evidence sources and L2G definitions followed Minikel *et al*. pQTL rows use a background restricted to proteomically measured targets, to control for panel-selection ascertainment, whereas non-pQTL rows use the full target–indication universe so as to reproduce the framework of Minikel et al; the two denominators are therefore not interchangeable. Neale UK Biobank, FinnGen and GWAS Catalog are Open Targets Genetics sub-sources sharing a common L2G gene assignment, and All OTG is their union; PICCOLO denotes eQTL colocalization-based gene assignment (Online Methods). Per-platform rows use platform-specific backgrounds, and three of the 23 launched pQTL-supported pairs are supported on both Olink and SomaScan, so per-platform counts do not sum to the overall pQTL row. **b**. Upset plot showing clinically successful T–I pairs often have multiple MR-coloc support types (cis, trans, mixed). **c**. Phase I– Launch RS by protein family (Minikel classification), comparing L2G share > 0.5 vs L2G share > 0.5 + pQTL evidence; genes could belong to multiple families. A[G] = genetically supported T–I pairs that launched (among those entering Phase I); S[G] = genetically supported T–I pairs entering Phase I; A![G] = genetically unsupported T–I pairs that launched (among those entering Phase I); S![G] = genetically unsupported T–I pairs entering Phase I. Error bars: 95% CI.

When we examined across MR-coloc subtypes, we found similar enrichment estimates with overlapping confidence intervals from mixed-, cis- or trans-MR, including a subset with cis-/trans-colocalization support (**Fig. 1a**). The similar RS across MR-coloc subtypes in our results and in previous studies motivated us to examine whether this pattern reflected overlapping evidence streams rather than independent predictive power. Indeed, most pQTL-supported launched T-I pairs were supported by two or more MR-coloc types, often combining cis, trans, and mixed MR with cis or trans colocalization, with intersection sizes up to four (**Fig. 1b, Supplementary Table 5**), highlighting the value of integrating evidence from multiple MR-coloc subtypes when prioritizing therapeutic hypotheses.

We also investigated whether enrichment was concentrated in certain druggable families using protein family annotations^11^. Under L2G alone, many well-known protein families like enzymes and kinases showed little or no enrichment, but when combined with pQTL support, they exhibited the largest RS increases (**Fig. 1c, Supplementary Table 6**). However, several medically important families (rhodopsin-class GPCRs, nuclear receptors, ion channels and many transporters) are poorly represented on current high-throughput Olink and Somascan panels, and in our data these classes have null enrichment; we suspect inadequate assay coverage (rather than absence of true biology) explains at least part of this gap. These results also imply that a straightforward way to boost genetic target validation is to expand and diversify proteomic panels by increasing coverage of existing panels for these under-represented classes or by employing novel methods to quantify low-abundance proteins and protein isoforms^12,13^.

We examined whether pQTL-supported enrichment varied by therapeutic area or clinical phase (**Supplementary Figures S2–S3 and Tables 7-15**). Although pQTL-supported target–indication pairs were unevenly distributed across therapeutic areas, enrichment estimates were broadly consistent across disease categories and phases, with some evidence of stronger enrichment for later-stage transitions. Notably, pQTL-supported target–indication pairs were not preferentially enriched in therapeutic areas with higher baseline success rates, arguing against confounding by disease category (**Supplementary Table 10-12**). Enrichment was homogeneous across therapeutic areas (Breslow–Day p = 0.21; **Supplementary Table 13**) and robust to leave-one-out exclusion of any single therapeutic area, with all lower confidence bounds remaining above 1 (**Supplementary Tables 14–15**).

A by-product of this work is a browsable database of target–trait pairs supported at a liberal pQTL MR FDR-corrected threshold (p < 0.05), available at https://mk31.shinyapps.io/pqtl_mr_fdr05/. Given the liberal threshold, false positive associations are expected; we therefore provide extensive downstream annotations to support follow-up and prioritization (**Online Methods**).

Using the combined evidence framework, we note several pQTL-based target-indication pairs, where the targets have a low or absent L2G score from Open Targets Genetics but are supported by other orthogonal sources of evidence (**Supplementary Table 16**). For example, genetically predicted higher tumor necrosis factor (TNF) was associated with increased risk of ankylosing spondylitis, consistent with the clinical efficacy of TNF inhibitors, yet this association is absent from Open Targets Genetics L2G predictions due to the exclusion of the complicated HLA/MHC region which is difficult to include in the post-GWAS analysis pipeline. Although this exclusion reduces confounding due to complex linkage disequilibrium, it can also remove informative associations from nearby genes such as *TNF*. We also observed non-HLA examples illustrating similar complementarity, for example *SOST* with osteoporosis, where MR evidence is supported by mechanistic data and approved therapies despite modest L2G scores in Open Targets Genetics^14,15^, highlighting the importance of MR as a complementary line of evidence for revealing likely causal genes for disease association.

We also point out associations, especially trans-associations, where therapeutic directionality inference in the absence of other supporting data can be misleading. For example, we noted a trans-colocalizing signal 17_39871710_C_G - near *CSF3* (colony stimulating factor 3) gene - which was associated with higher plasma CSF3R (colony stimulating factor 3 receptor) levels and higher risk of neutropenia inconsistent with the function of CSF3R which typically increases granulocyte counts^16^. The CSF3R-neutropenia relationship was, however, consistent with CSF3R function when considering cis-acting variants (i.e. variants near the *CSF3R* gene). This highlights the need to consider therapeutic direction inferred from trans-acting variants in context of their cis-acting counterparts since trans-signals may represent complex biological feedback mechanisms and, additionally, plasma protein levels may not reflect the cellular or tissue-specific context in which a target exerts its therapeutic effect.

There are several limitations to our work. First, the 23 pQTL-supported and launched T-I pairs supporting the current RS estimates are driven by only 12 targets, reflecting the strong dependence of proteogenomic validation on protein assay coverage; broader and more diverse protein panels are likely to expand this set. Second, some cis pQTL associations may be influenced by protein-altering variants that affect aptamer- or antibody-based measurements, potentially complicating direction-of-effect interpretation, although we annotate such variants to support cautious follow-up (**Supplementary Table 16**). Third, the underlying MR and colocalization analyses rely primarily on common variants from European-ancestry cohorts and on circulating plasma proteins, which may not reflect disease-relevant tissues and limit generalizability. Finally, we did not assess eQTL-based enrichment, as most large-scale eQTL resources are used in cis and are not directly comparable to the genome-wide pQTL framework used here. Our analyses are also largely naive to the direction of genetic effect, which is unknown or unannotated in most of the datasets used, so a fully direction-aware relative success estimate was not feasible; among launched pQTL-supported pairs, 17 of 23 were directionally aligned with the mechanism of the launched agent, and excluding non-aligned pairs from the numerator gave a relative success of 3.28 (95% CI: 2.22, 4.84) (**Supplementary Tables 17–18**). The confidence intervals reported here are conditional on the observed set of 12 contributing targets; intervals computed with a cluster-robust variance clustered on target gene are provided in **Supplementary Table 21**. Launched pQTL-supported pairs are concentrated at high L2G share, with 23 of 26 at 0.75 or above, so the standalone contribution of pQTL evidence at low L2G share is not estimable from these data (**Supplementary Table 22**). Finally, we acknowledge that a matched genome-wide eQTL Mendelian randomization resource, spanning cis and trans effects across the same breadth of traits, might recover part of the enrichment we attribute to pQTLs; separating resource breadth from molecular layer will require such a resource.

### Online Methods

We used eight publicly accessible proteogenomic datasets^10,17–23^ (**Supplementary Table 1)** for the pan-MR analyses using GSMR enabling selection of genetic instruments from across the genome; regional definitions were applied post-GSMR, i.e. cis-, defined as +1Mbp from transcription start site, trans-, or mixed-MR. The MR associations were expressed per standard deviation (SD) higher genetically predicted plasma protein concentrations. A subset of MR associations (FDR-corrected p-value < 0.05) also underwent genetic colocalization tests. Colocalization support was defined as H4 ≥ 0.8, recorded separately for cis and trans loci. We combined complex trait GWAS selected as outcomes from our previous release^9^ with new traits from FinnGen (release 12), pan-UK Biobank, and GWAS Catalog, using the same criteria we used previously, i.e. - if they had at least one genome-wide significant (p < 5 x 10^-8^) locus, ensuring a systematic disease-agnostic trait selection strategy. This resulted in a total 8762 studies (**Supplementary Table 2)**. A list of studies that were removed due to their lack of medical relevance or redundant protein associations is provided in **Supplementary Table 3**. Details of genotyping protocols and QC of proteomic studies have been described previously in the respective studies’ publications. Details of liftOver and harmonization of both proteomic and outcome studies are provided in our previous preprint^9^.

For comparability of pQTL-based enrichment estimates with other sources of genetic evidence, we followed Minikel et al’s approach. We linked targets to indications using the MeSH term based similarity matrix and kept target-indication (T-I) pairs with a similarity score ≥ 0.8. For associations from Open Targets Genetics (version 8), including the traits used in our pQTL MR analyses, we additionally required an L2G share of at least 0.5 (L2G share is the L2G score of a gene divided by the sum of L2G scores across all candidate genes for that trait–loci combination from Open Targets Genetics^24^). A target–indication (T-I) pair was considered pQTL-supported if the MR p-value passed a Bonferroni threshold (p ≤ 0.05 / 4.7×10^7^, ∼1.06×10^−9^) and the pair satisfied the MeSH and L2G filters. A stricter “MR+coloc” group also required a cis- or trans-colocalization posterior probability H_4_ ≥ 0.8. For each T-I pair we selected the row with the highest clinical phase and, if needed, breaking ties by the MeSH similarity score. Using this set, we computed Phase I to launch relative success as RS = (A[G]/S[G]) / (A[!G]/S[!G]), where S is the number of Phase I entries and A the number of launches in the genetically supported (G) and unsupported (!G) groups. Confidence intervals for RS were obtained using the Katz log method, and where p-values are reported, they were calculated using Fisher’s exact test. For pQTL-based analyses, we restricted the background set to Phase I T–I pairs whose target protein was measured on proteomic platforms of the respective pQTL studies (e.g. Olink). For other genetic evidence sources, the background consisted of Phase I T-I pairs that lacked that specific source of support as defined by Minikel et al. PICCOLO support was taken as defined by Minikel et al: putative effector genes assigned by colocalization between GWAS Catalog associations (p < 5 × 10^−8^) and molecular QTLs from GTEx and other published resources spanning 68 tissues and cell types, using the PICS algorithm with reference linkage-disequilibrium values in place of full summary statistics^25^. We retained PICCOLO-supported pairs at an eQTL p < 1 × 10^−5^ and colocalization posterior probability H_4_ ≥ 0.9. Effector genes for all Open Targets Genetics sub-sources shown in Figure 1a — Neale UK Biobank, FinnGen and GWAS Catalog — were assigned by the same L2G model, so these labels denote study provenance within a single Open Targets Genetics release rather than separate gene-assignment procedures; associations were required to reach p < 5 × 10^−8^. Definitions, thresholds and gene assignments for all non-pQTL evidence sources, including OMIM, Genebass and PICCOLO, were inherited unchanged from Minikel et al and were not recomputed here. Support therefore required a Bonferroni-significant association with the MeSH-matched indication rather than the mere existence of a pQTL for the target (**Supplementary Table 20**). Confidence intervals for relative success are Katz intervals on a 2 × 2 table, following Minikel et al, and are reported without adjustment for within-target correlation so that they remain comparable with the L2G-based estimates reproduced in Figure 1a; intervals computed with a cluster-robust variance clustered on target gene, across all 753 contributing targets, are provided in **Supplementary Table 21**.

To assess whether pQTL-based enrichment was confounded by the therapeutic area, we performed several sensitivity analyses. First, we tested whether pQTL evidence was disproportionately distributed across therapeutic areas using a chi-square test (**Supplementary Table 7**). Second, we evaluated whether pQTL support was preferentially found in therapeutic areas with high baseline success rates using Spearman rank correlation between each therapeutic area’s baseline success rate (among pQTL-unsupported targets) and the proportion of target-indication pairs with pQTL support (**Supplementary Table 10**). Third, we applied the Breslow-Day test for homogeneity of odds ratios across therapeutic area strata to assess whether the pQTL effect was consistent across disease categories (**Supplementary Table 13**). These tests were performed using two background universes: (1) all Phase I-entered target-indication pairs where the gene was measurable on proteomic platforms (Olink/SomaScan), and (2) the subset of these with existing genetic evidence from Open Targets Genetics, OMIM, PICCOLO, or Genebass (**Supplementary Tables 8-9, 11-12, 14-15**). As an additional sensitivity analysis, we also computed pQTL enrichment using the full Minikel et al. T-I universe without restricting to measured proteins (**Supplementary Figure S4**). Finally, we performed leave-one-therapeutic-area-out analyses to evaluate whether overall RS estimates were sensitive to removal of any single disease category (**Supplementary Tables 14-15**).

To assess whether cis-association signals were protein-altering variants (PAVs) or in linkage disequilibrium with PAVs, we used the TOP-LD tool^26^. Starting from Bonferroni-significant cis-MR index SNPs, we queried TOP-LD in Europeans (r^2^ ≥ 0.6, MAF ≥ 0.01) and extracted linked variants, their annotated genes, and Sequence Ontology (SO) consequences. We classified a variant as protein-altering if it carried a HIGH or MODERATE impact SO term^27^. For trans-associations, we recorded the nearest gene(s) for each trans index SNP from Open Targets. We also annotated replication across proteomic platforms and outcome datasets, the different combinations of MR and colocalization methods supporting each target-trait pair, and non-genetic evidence sources (for example, RNA expression, animal models, and literature) using Open Targets Platform^28^ (https://platform.opentargets.org/, version 25.09.0) data types. Finally, we overlaid Pharmaprojects and ChEMBL to flag known drug targets, positive control T-I pairs (existing programmes for the same indication), and potential repositioning opportunities (known targets without programmes for the MR-inferred indication). Further implementation details, including data processing, harmonisation, and all thresholds used in enrichment and annotation analyses, are provided in the **Supplementary Methods**. All analyses and annotations, unless otherwise specified, were done in R version 4.3.3. Therapeutic direction was assigned from the launched agent at the target–indication level. The expected direction of the cis effect was derived from analyte class (secreted effector, shed receptor ectodomain, or enzyme, assigned by target class), the direction of the drug, and the orientation of the matched GWAS trait, and this classification was fixed before comparison with the observed effect directions. Trait orientation was resolved against the matched GWAS trait rather than the MeSH indication term.

## Acknowledgements

We would like to thank Eric Vallabh Minikel for reviewing preliminary analyses and figures related to this work.

## Data availability

Summary data for both proteins and outcomes used for genetic analyses are publicly available from the GWAS catalog https://www.ebi.ac.uk/gwas/downloads/summary-statistics. Data on clinical precedence for approved and investigational drugs and representative target-indication pairs with L2G share, were obtained from Minikel et al. (2024) https://github.com/ericminikel/genetic_support/. An analytical MR and genetic colocalization dataset sufficient to reproduce all figures and statistics herein can be downloaded here: https://doi.org/10.5281/zenodo.18451758. Open Targets Genetics L2G data version 8 (22.02.01) is available from the EMBL-EBI FTP server: https://ftp.ebi.ac.uk/pub/databases/opentargets/genetics/. Filtered results (MR FDR-corrected p-value < 0.05) can be browsed here: https://mk31.shinyapps.io/pqtl_mr_fdr05/.

## Code availability

Source code to reproduce all figures and statistics in the present analysis is provided at https://github.com/mohdkarim/mrcoloc. Codes that were used to perform Mendelian randomisation are provided here.https://github.com/opentargets/mendelian-randomisation.

## Author contributions

MAK, MG, and JM conceived the study. MAK, AH, and BA collaboratively performed all analyses. MG supervised the research. JH and AB worked on curation, integration and harmonisation of the datasets for the GWAS Catalog. MAK and MG drafted the manuscript. All authors provided valuable feedback and critical comments that informed the design, analyses and interpretation of the study and reviewed and approved the final manuscript.

## Ethics declarations

All institutions contributing cohorts to the proteomics and outcome studies received ethics approval from their respective research ethics review boards.

---

**Supplementary Figure S1.**
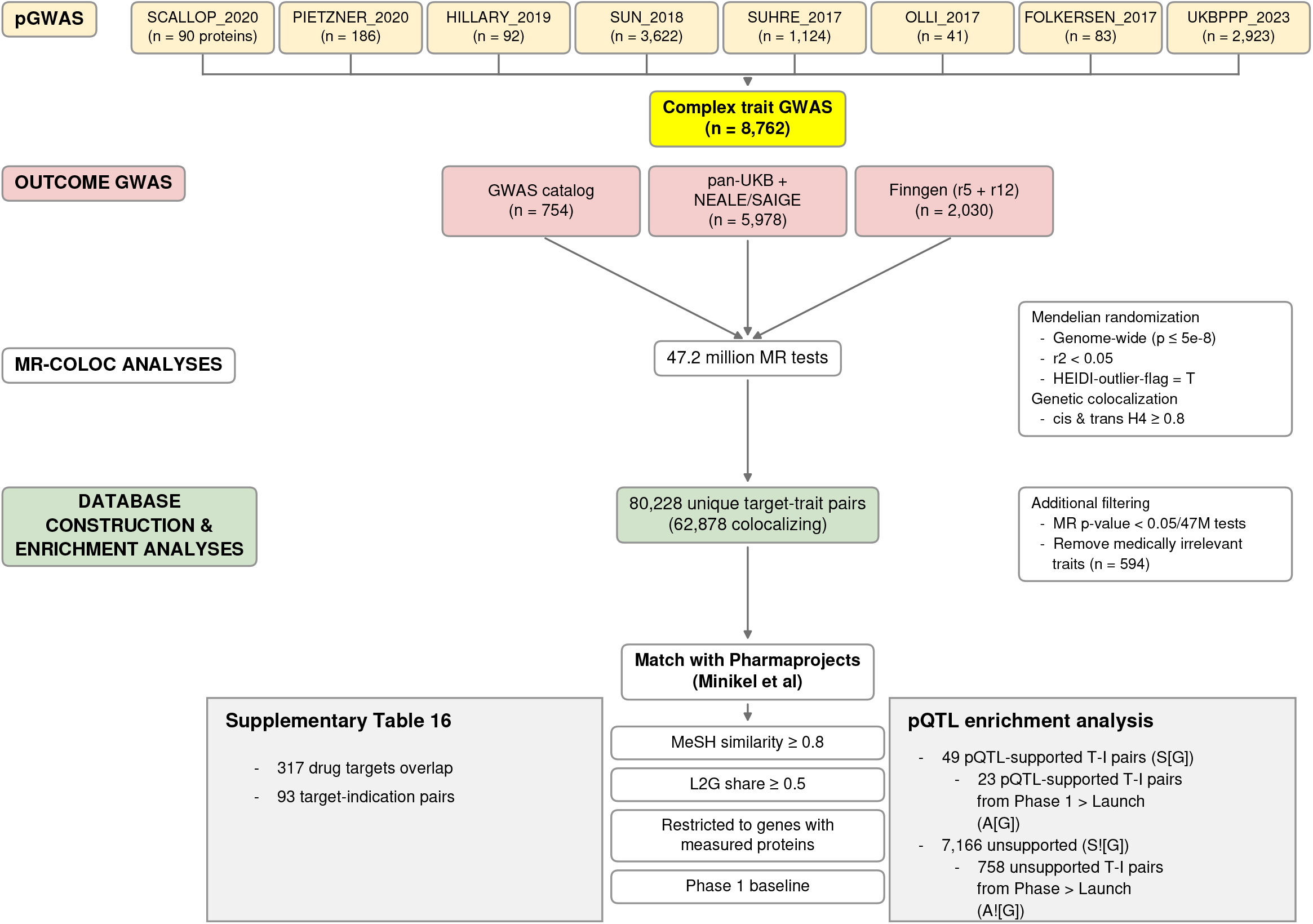
Study design flowchart illustrating the two analytical paths: (1) systematic identification of pQTL-supported target-trait pairs through Mendelian randomization of eight proteogenomic datasets against ∼8,000 complex trait GWAS, and (2) enrichment analysis comparing clinical success rates of pQTL-supported versus unsupported target-indication pairs using the Minikel et al. framework. Numbers at each step indicate the count of datasets, traits, associations, or T-I pairs retained after applying the indicated filters.

**Supplementary Figure S2.**
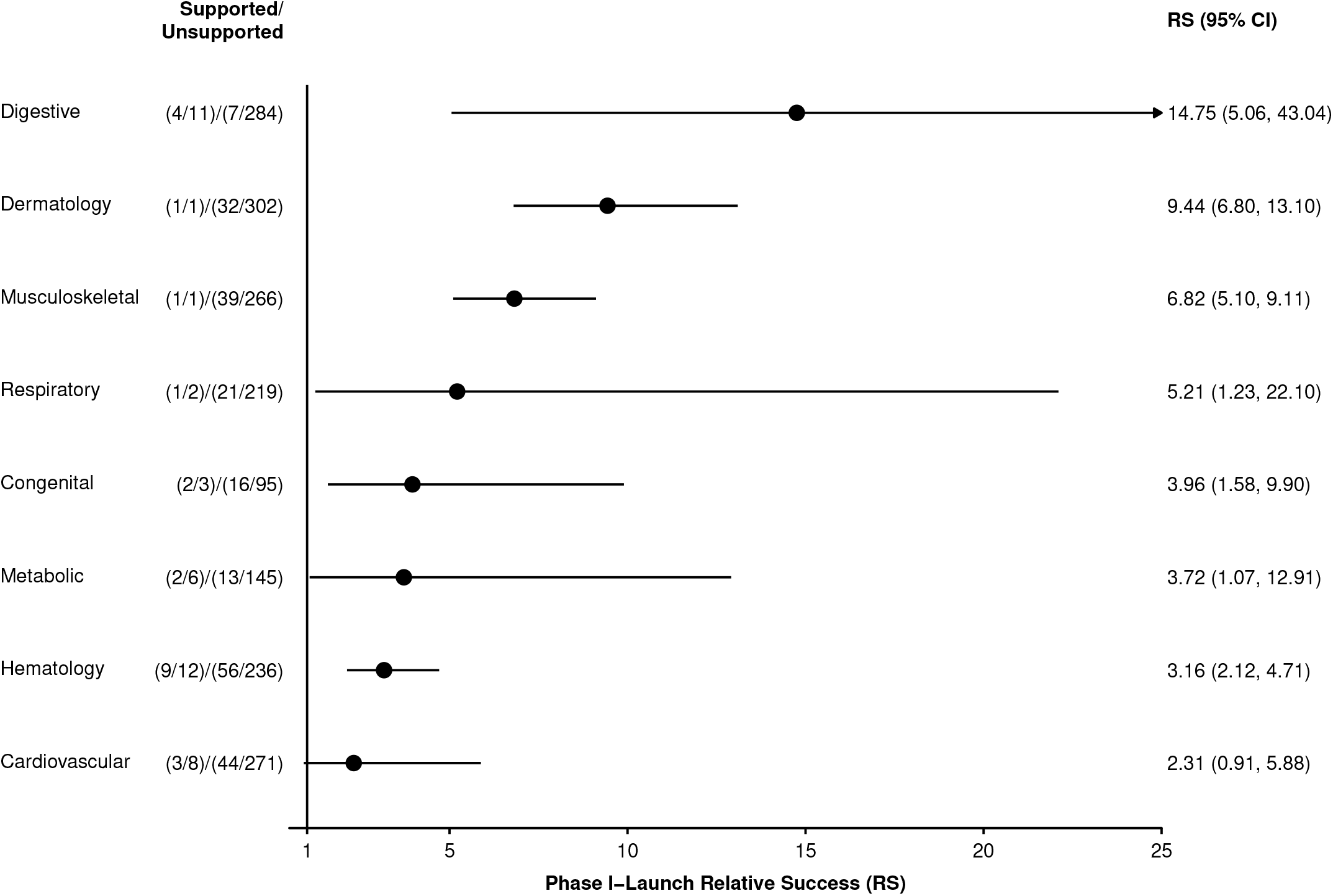
Phase I-Launch relative success (RS) of pQTL-supported versus unsupported target-indication (T-I) pairs by therapeutic area. RS was computed within each therapeutic area using the Katz log method. Therapeutic areas were included if they had at least one pQTL-supported T-I pair entering Phase I and at least one pQTL-supported pair that reached launch (RS > 0). Nine therapeutic areas were excluded: six with no pQTL-supported T-I pairs at Phase I (endocrine, immune, infection, ophthalmology, other, psychiatry) and three with pQTL-supported pairs but no launches, yielding RS = 0 (neurology, oncology, signs/symptoms). The background was restricted to T-I pairs where the target was measured on at least one proteomic platform (Olink or SomaScan). Supported = A[G]/S[G] = launched/Phase I among pQTL-supported pairs; Unsupported = A[!G]/S[!G] = launched/Phase I among pQTL-unsupported pairs. Error bars: 95% CI (Katz log method).

**Supplementary Figure S3.**
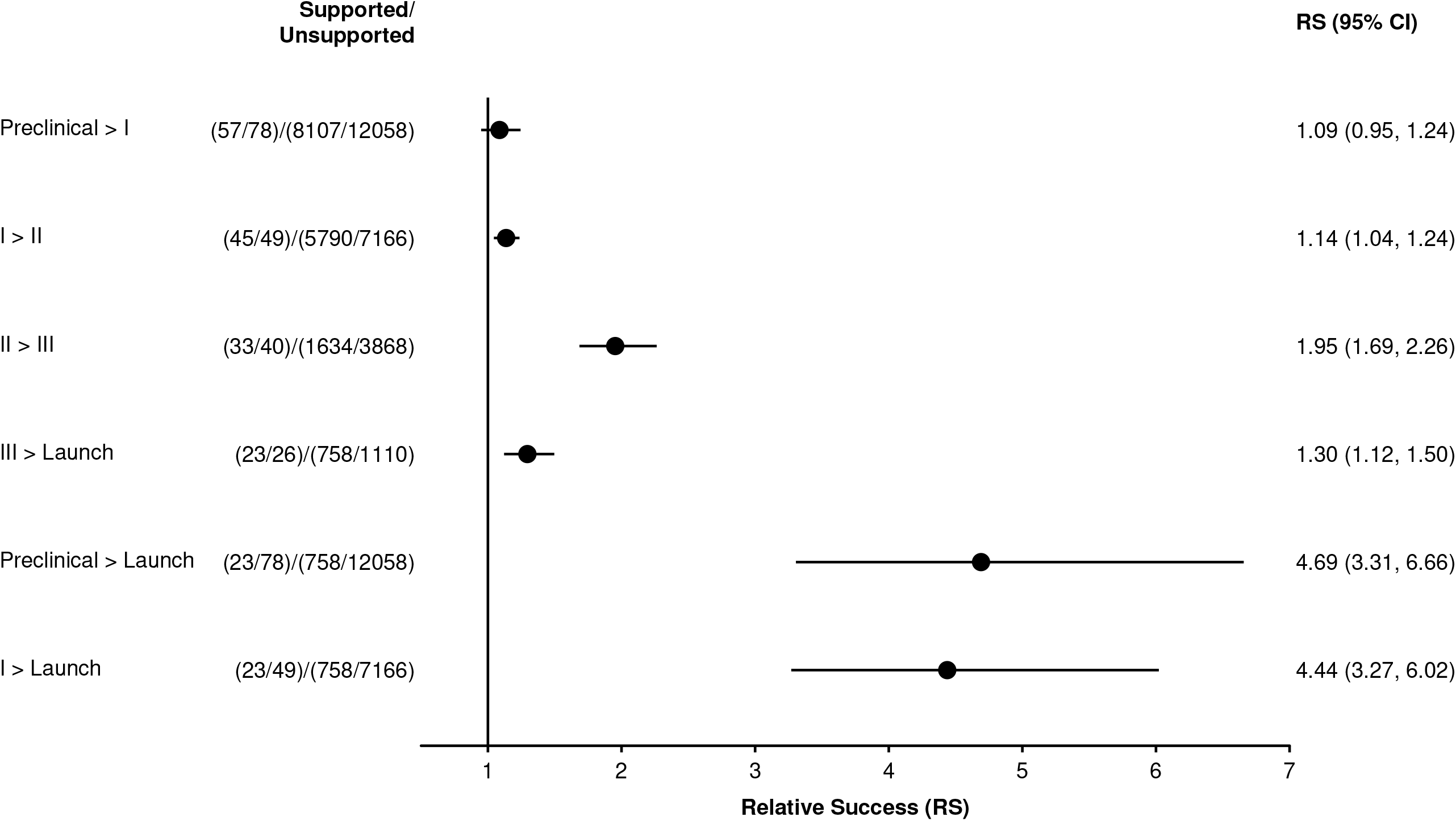
Relative success (RS) of pQTL-supported versus unsupported target-indication (T-I) pairs by clinical phase transition. RS was computed separately for each individual phase transition (Preclinical > I, I > II, II > III, III > Launch) and for cumulative transitions (Preclinical > Launch, I > Launch). The denominator for each transition includes only T-I pairs that entered the starting phase. The background was restricted to T-I pairs where the target was measured on at least one proteomic platform (Olink or SomaScan). Supported = A[G]/S[G] = succeeded/entered among pQTL-supported pairs; Unsupported = A[!G]/S[!G] = succeeded/entered among pQTL-unsupported pairs. Error bars: 95% CI (Katz log method).

**Supplementary Figure S4.**
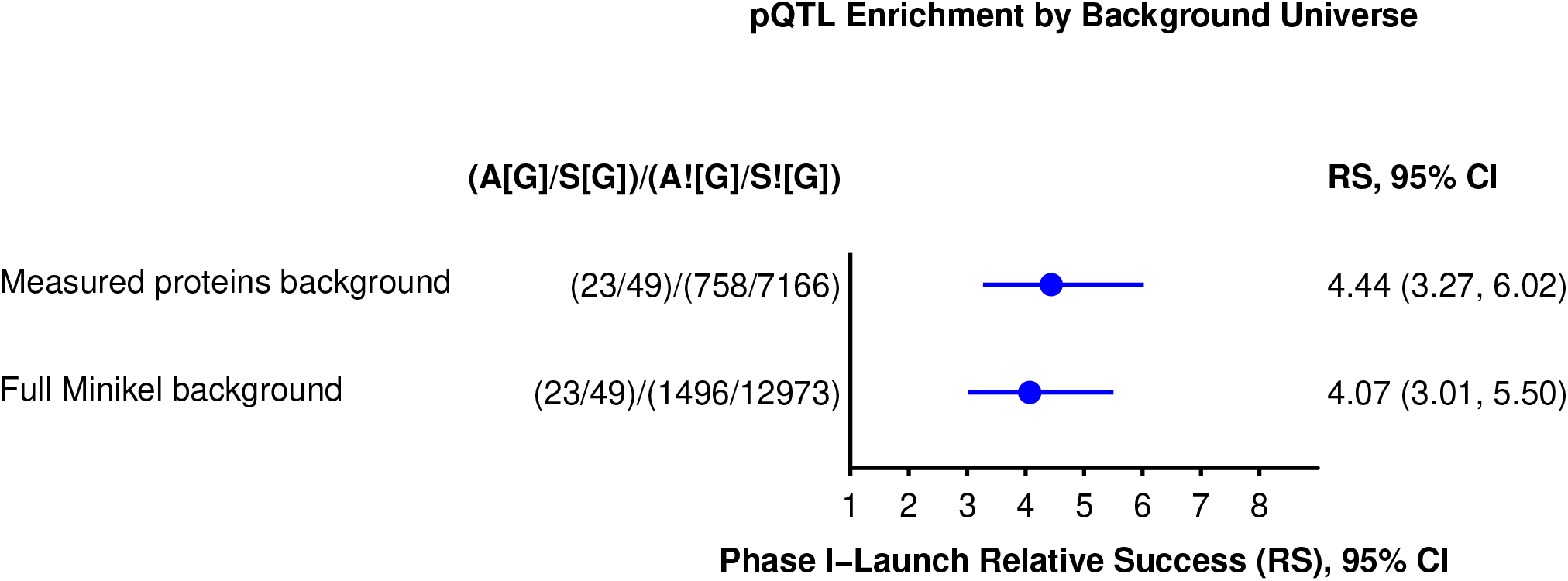
Sensitivity analysis comparing Phase I-Launch relative success (RS) of pQTL-supported T-I pairs under two background definitions: (1) measured proteins background, restricted to T-I pairs where the target was assayed on at least one proteomic platform used in the pQTL studies (Olink or SomaScan), matching the main analysis; and (2) full Minikel background, using all T-I pairs from Minikel et al. without restriction to measured proteins. pQTL-supported T-I pairs are identical in both analyses; only the unsupported comparator group changes. A[G]/S[G] = launched/Phase I among pQTL-supported pairs; A[!G]/S[!G] = launched/Phase I among pQTL-unsupported pairs. Error bars: 95% CI (Katz log method).

## References

1. Minikel, E. V., Painter, J. L., Dong, C. C. & Nelson, M. R. Refining the impact of genetic evidence on clinical success. Nature 629, 624–629 (2024).

2. King, E. A., Davis, J. W. & Degner, J. F. Are drug targets with genetic support twice as likely to be approved? Revised estimates of the impact of genetic support for drug mechanisms on the probability of drug approval. PLoS Genet 15, e1008489 (2019).

3. Nelson, M. R. et al. The support of human genetic evidence for approved drug indications. Nature Genetics 47, 856–860 (2015).

4. Mountjoy, E. et al. An open approach to systematically prioritize causal variants and genes at all published human GWAS trait-associated loci. Nat Genet 53, 1527–1533 (2021).

5. Deng, Y.-T. et al. Atlas of the plasma proteome in health and disease in 53,026 adults. Cell 188, 253–271.e7 (2025).

6. Zheng, J. et al. Phenome-wide Mendelian randomization mapping the influence of the plasma proteome on complex diseases. Nat Genet 52, 1122–1131 (2020).

7. Ferolito, B. R. et al. Leveraging Large-Scale Biobanks for Therapeutic Target Discovery. medRxiv 2025.02.10.25321487 (2025) doi:10.1101/2025.02.10.25321487.

8. Duffy, Á. et al. Development of a human genetics-guided priority score for 19,365 genes and 399 drug indications. Nat Genet 56, 51–59 (2024).

9. Karim, M. A. et al. Systematic disease-agnostic identification of therapeutically actionable targets using the genetics of human plasma proteins. medRxiv 2023.06.01.23290252 (2023) doi:10.1101/2023.06.01.23290252.

10. Sun, B. B. et al. Genomic atlas of the human plasma proteome. Nature 558, 73–79 (2018).

11. Minikel, E. V. et al. Evaluating drug targets through human loss-of-function genetic variation. Nature 581, (2020).

12. Lu, C., Bonini, A., Viel, J. H. & Maglia, G. Toward single-molecule protein sequencing using nanopores. Nat Biotechnol 43, 312–322 (2025).

13. Motone, K. et al. Multi-pass, single-molecule nanopore reading of long protein strands. Nature 633, 662–669 (2024).

14. Döring, Y. et al. CXCL12 Derived From Endothelial Cells Promotes Atherosclerosis to Drive Coronary Artery Disease. Circulation 139, 1338–1340 (2019).

15. Aditya, S. & Rattan, A. Sclerostin Inhibition: A Novel Target for the Treatment of Postmenopausal Osteoporosis. J Midlife Health 12, 267–275 (2021).

16. Dwivedi, P. & Greis, K. D. Granulocyte colony-stimulating factor receptor signaling in severe congenital neutropenia, chronic neutrophilic leukemia, and related malignancies. Exp Hematol 46, 9–20 (2017).

17. Folkersen, L. et al. Mapping of 79 loci for 83 plasma protein biomarkers in cardiovascular disease. PLoS Genet 13, e1006706 (2017).

18. Folkersen, L. et al. Genomic and drug target evaluation of 90 cardiovascular proteins in 30,931 individuals. Nat Metab 2, 1135–1148 (2020).

19. Hillary, R. F. et al. Genome and epigenome wide studies of neurological protein biomarkers in the Lothian Birth Cohort 1936. Nat Commun 10, 3160 (2019).

20. Pietzner, M. et al. Genetic architecture of host proteins involved in SARS-CoV-2 infection. Nat Commun 11, 6397 (2020).

21. Suhre, K. et al. Connecting genetic risk to disease end points through the human blood plasma proteome. Nat Commun 8, 14357 (2017).

22. Ahola-Olli, A. V. et al. Genome-wide Association Study Identifies 27 Loci Influencing Concentrations of Circulating Cytokines and Growth Factors. Am J Hum Genet 100, 40–50 (2017).

23. Sun, B. B. et al. Plasma proteomic associations with genetics and health in the UK Biobank. Nature 622, (2023).

24. Ghoussaini, M. et al. Open Targets Genetics: systematic identification of trait-associated genes using large-scale genetics and functional genomics. Nucleic Acids Res 49, D1311–D1320 (2021).

25. Guo, C. et al. Identification of putative effector genes across the GWAS Catalog using molecular quantitative trait loci from 68 tissues and cell types. bioRxiv (2019) doi:10.1101/808444.

26. Huang, L. et al. TOP-LD: A tool to explore linkage disequilibrium with TOPMed whole-genome sequence data. Am J Hum Genet 109, 1175–1181 (2022).

27. McLaren, W. et al. The Ensembl Variant Effect Predictor. Genome Biology 17, 122 (2016).

28. Buniello, A. et al. Open Targets Platform: facilitating therapeutic hypotheses building in drug discovery. Nucleic Acids Res 53, D1467–D1475 (2025).

